# Design and Formative Evaluation of a Voice-based Virtual Coach for Problem-Solving Treatment

**DOI:** 10.1101/2021.05.13.21257041

**Authors:** Thomas Kannampallil, Corina R. Ronneberg, Nancy Wittels, Vikas Kumar, Nan Lv, Joshua M. Smyth, Ben S. Gerber, Emily Kringle, Jillian A. Johnson, Philip Yu, Lesley E. Steinman, Olu Ajilore, Jun Ma

## Abstract

Artificial intelligence (AI)-based voice technology offers considerable promise in healthcare; however, its application for behavioral therapy in a real-world or research setting has yet to be determined. We describe the design and evaluation of ^1^Lumen™, a fully functional voice-only virtual coach that delivers evidence-based, problem-solving treatment (PST) for patients with mild-to-moderate depression and/or anxiety. Participants (N=26) completed surveys and semi-structured interviews after two therapy sessions with Lumen. Participants found Lumen to provide high pragmatic usability and favorable user experience, with marginal task load during interactions. Participants emphasized its on-demand accessibility and the delivery of a complex PST task with a simplistic structure for achieving therapy goals. Suggestions for improvement included streamlining and improved personalization and pacing of conversations and providing additional context during therapy sessions. Lumen offers a realistic and cognitively plausible verbal interaction that can potentially lead to personalized and accessible mental health care, filling a gap in traditional mental health services.

## Introduction

Artificial Intelligence (AI) has afforded new opportunities for human interactions with technology for the practice of medicine [1]. Of the recent innovations, voice assistants that rely on AI-based platforms such as Amazon’s Alexa, Google Home, Cortana and Siri, have transformed how humans search for information, with recent reports suggesting that nearly 30% of search queries use voice-based input [2]. Broad adoption of such platforms lends support for their potential utility in healthcare-related applications such as behavioral counseling to promote healthy lifestyle habits and emotional well-being [3, 4]. However, current healthcare-related applications of voice assistants are generally rudimentary and few have been developed for delivering evidence-based therapies or have been subjected to careful evaluation (e.g., to inform development, or for their effect on clinical or behavioral outcomes) [5]. Towards this end, we developed and evaluated, Lumen™, an end-to- voice-only virtual coach that delivers evidence-based, problem-solving treatment (PST) for patients with mild-to-moderate symptoms of depression and/or anxiety.

Lumen, by design, is different from the current spectrum of voice assistant health applications that primarily support information seeking activities [4]. Studies on information seeking activities performed on voice assistants have focused on the quality and content of voice assistant responses for myriad topics including health behavior and lifestyle [6, 7], mental health, interpersonal violence, and addiction help [8, 9], patient and consumer safety risks [10], vaccines [11], medication names [12], and sexual health [13]. The findings across these studies consistently highlighted the challenges associated with information seeking using voice-based search in medical/care contexts. For example, Bickmore et al. found that the Siri, Alexa and Google Assistant platforms (and their underlying algorithms) were effective in completing only 43% of requests regarding situations that required medical expertise, and 29% of the responses could have resulted in some degree of patient harm [10]. Fewer applications for assessment and support have been developed; these applications have generally been preliminary or simple prototypes that have been used for delivering visual acuity test [14], support for coping with chronic disease [15], and for nutritional planning [16]. These applications have largely lacked outcome assessment or incorporation of behavioral therapy [4].

In this paper, we describe the design and formative evaluation of Lumen, focusing on (a) the user experience, task-related workload associated with interactive communication, and participant-alliance with delivered treatment; and (b) user perspectives including the benefits, challenges and barriers to Lumen use, and recommendations for design improvements. This formative evaluation is a precursor to an on-going randomized mechanistic trial (ClinicalTrials.gov, NCT# 04524104) for investigating the treatment efficacy of using Lumen as a virtual coach for treating mild-to-moderate depression and anxiety.

## Method

In the following sections, we describe the design components of Lumen, its features, and the mixed-method study that was conducted.

### Lumen

Lumen is a voice-only virtual coach that delivers an evidence-based, 8-session PST program for patients with mild-to-moderate depression and/or anxiety. Lumen’s design was based on two overarching principles: (a) providing cognitively plausible conversations, i.e., aligning Lumen’s conversations with the cognitive processes of human communicative interactions [5], and (b) alignment with the principles of evidence-based PST. This PST program was previously tested and delivered with a human coach [17]; Lumen incorporates essential components of that treatment protocol for coaching and monitors progress using surveys and ecological momentary assessments (EMAs). All of the Lumen design components are delivered in an integrated environment, coordinated through the voice-only platform and associated mobile tools.

Developed on Amazon’s Alexa platform, the Lumen architecture incorporates an intelligent conversation manager that manages the content, structure and flow of interactive conversations between a patient and Lumen, and a context manager that incorporates context awareness to the conversations. Utilizing underlying AI capabilities of the Alexa platform, the conversation manager utilizes user verbal input (“intents”) to provide appropriate, synchronous responses, aligned with PST’s treatment guidelines. The PST content and conversational structure were designed in consultation with master PST trainers and PST experts. The context manager provides contextual awareness to the interactions by incorporating user input from surveys and EMAs (delivered asynchronously through mobile applications) and treatment progression and continuity (e.g., review of patient problems and action plans from a previous session) (Details of the Lumen architecture and features are provided in Supplementary Material, sections A, B).

We followed an iterative user-centered design process, comprised of brainstorming sessions with software engineers, interaction designers, psychiatrists, and researchers; prototype development on the Alexa platform; and several iterations of internal testing.

### Participants and Study Design

Participants for this formative evaluation were recruited from the recently completed, ENGAGE-2 trial (ClinicalTrials.gov, NCT# 03841682), in which a PST-certified health coach delivered integrated collaborative care for depression and obesity to intervention participants, whereas those in the control group received usual care. A convenience sample (n=91 of 106) of ENGAGE-2 participants were contacted for assessing their interest in participating in a study with a virtual PST coach. Of these participants, 26 expressed interest and consented to participate: 17 had prior PST experience (i.e., part of the ENGAGE-2 intervention group) and 9 did not (i.e., part of the ENGAGE-2 control group). The study was approved by the institutional review board of the University of Illinois (IRB#2020-0918).

This was an observational study, with each participant completing two Lumen sessions: an *introductory* first session (termed S1; n=26) and a *problem-solving* second session (termed S2; n=24, missing 2 of the 9 ENGAGE-2 control participants). The two sessions were representative of the 8-session, evidence-based PST evaluated in a previous trial [17]. During S1, Lumen provides a program overview, a detailed introduction to the PST process and behavioral activation and guides the participant to create a list of problems to address in subsequent sessions. In S2, Lumen guides the participant through the steps of problem-solving: identifying a problem to address, setting a goal, brainstorming possible solutions, evaluating the pros and cons of each solution, selecting a solution to implement, and developing an action plan to carry out before the next session. S2 concludes with behavioral activation coaching, where Lumen assists participants with selecting a social, physical, and pleasant activity to partake in prior to the next session. The full Lumen PST program will include six more problem-solving sessions that follow the same structure as S2, which was the rationale for testing only one problem-solving session during this formative evaluation.

The purpose of the two-session approach was to conduct a representative evaluation of all Lumen sessions and to evaluate whether there were differences in participant experience and interactions between the sessions.

### Procedure

Consented participants were provided access to the Lumen S1 and S2 skills via the Alexa application and were given instructions on how to enable the skills on the Alexa app on their personal phones or mobile devices. All user interviews were conducted remotely by a trained research coordinator and note taker via a video conferencing system (Zoom). Participants were first provided with a brief overview of the study purpose and their access to the Lumen skill (designed as a private skill, that was available by invitation only) was verified. The research coordinator went through a list of tips for effectively communicating with the Lumen coach and answered any questions that arose. After this, participants were instructed to turn off their video, and audio recording via Zoom was enabled from this point. Participants then opened the Alexa app, and said “Open Lumen Coach” to begin their Lumen session. During their Lumen sessions, the trained note taker took notes of any deviations from the session script or any technical problems.

After each Lumen session (S1 and S2), the coordinator followed a semi-structured interview script that included the following components: first, participants were asked to walkthrough their interaction experience with Lumen during their completed session, reflecting on what worked, what did not, and challenges they faced. Although the same procedure was followed for both Lumen sessions, interview questions varied slightly from S1 to S2 in order to inquire about session specific content. Interview questions after S1 focused on participant impressions of Lumen, suggestions for improving Lumen, evaluating the usefulness of tips around how to communicate with Lumen, and impressions of the PST overview. Interview questions after S2 included questions about participant impressions of Lumen that were different from S1, delivery of PST by Lumen, and factors affecting their likelihood of Lumen use in the future.

After interviews concluded, participants were emailed a link to 3 brief post-interview surveys related to user experience, workload, and the collaborative relationship between the participant and Lumen (User Experience Questionnaire Short Version (UEQ-S) [18], NASA Task Load Index (TLX) [19], and Working Alliance Inventory–Technology Version (WAI-Tech) [20]).

Audio recordings of interviews (26 for S1, 24 for S2) were transcribed with Trint audio transcription software for subsequent analysis. A total of 26 post-interview surveys (100%) were completed after S1, and 23 after S2 (95%).

### Data Analysis

Data analysis included coding of interview transcripts using thematic analyses and descriptive summaries of the user experience, task load, and WAI-Tech surveys.

### Coding of Transcripts

All interview transcripts were coded using an inductive thematic analysis to characterize the participant perspectives regarding their interaction with Lumen [21]. This approach involved the following stages: first, two co-authors (CR, EK) read the interview transcripts to familiarize with the content. Next, a set of “open codes” were created to characterize the content and context discussed in the interviews (i.e., inductive coding) [22]. These initial codes were compared across the transcripts to identify repeated and interrelated sub-themes. Similar sub-themes were grouped over multiple review sessions to develop a set of six overarching themes. Two co-authors (EK, CR) independently coded a set of 5 transcripts with a high degree of inter-rater agreement (Cohen’s Kappa ranged from 0.83-1 with a mean of 0.93). Discrepancies were resolved through discussion with the first author (TK). Following this, all remaining transcripts were coded.

### Surveys

From the UEQ-S survey, pragmatic quality and hedonic quality scale values were calculated by rescaling the survey responses to the range -3 to 3 and calculating item means within each scale using the UEQ-S Data Analysis Tool [23]. Pragmatic quality refers to the task or goal-related interaction qualities (e.g., efficiency, perspicuity, dependability) that a user aims to reach when using the product. Hedonic quality refers to the aspects related to pleasure or fun (e.g., stimulation, novelty) while using the product. Values <-0.8 represent a negative evaluation, between -0.8 and 0.8 represent a neutral evaluation, and >0.8 represent a positive evaluation on each scale.

The NASA TLX rating sheet was administered assuming similar weights for each of the 5 task load items (except for physical demand which was irrelevant to Lumen): mental demand, temporal demand (e.g., being rushed), effort, frustration and performance. Each item was then rescaled to the range 5 to 100 by multiplying raw score by 5.

From the WAI-Tech survey, three 12-item subscales (task, goal, bond) scores and an overall score were calculated as item means within each subscale. The task subscale reflected how responsive Lumen was to the participant’s focus or need, the goal subscale reflected the extent to which goals were important, mutual, and capable of being accomplished, and the bond subscale reflected the degree of mutual liking and attachment [20]. A higher overall score reflected a more positive rating of working alliance.

Given that the two sessions focused on two primary components of PST sessions—a session overview, and a problem-solving session—we evaluated whether there were differences in the user experience, task load or work alliance between these sessions. Towards this end, scores on each of the scales between S1 and S2 were compared using paired *t* tests. Analyses were conducted using SAS, version 9.4 (SAS Institute Inc., Cary, North Carolina); statistical significance defined by 2-sided P<0.05.

## Results

### General Characteristics

Among the 26 participants, 76.9% (n=20) were female and 73.1% were racial/ethnic minorities (50.0% Black, 23.1% Hispanic) with an average age of 43.9 years (SD, 11.9 years); 38.5% had a high school or some college education, and 53.8% had an annual family income <$55,000 (Table 1). Participants with previous PST experience (n=17) and those without (n=9) did not differ in age, race, income or educational status, although 11 of the 17 vs. 9 of the 9 were female (*P*=0.04).

**Table 1.** Baseline Characteristics by Prior PST Experience.

| Characteristic | All Lumen<br>formative<br>evaluation<br>participants<br>(n=26) | With prior PST<br>experience<br>(n=17) | Without prior<br>PST experience<br>(n=9) | P<br>value |
| --- | --- | --- | --- | --- |
| Age, years, mean ± SD | 43.9 ± 11.9 | 42.6 ± 13.2 | 46.3 ± 9.2 | 0.46 |
| Female, n (%) | 20 (76.9) | 11 (64.7) | 9 (100.0) | 0.04 |
| Race/Ethnicity, n (%) |  |  |  | 0.34 |
| Non-Hispanic White | 4 (15.4) | 3 (17.7) | 1 (11.1) |  |
| African American | 13 (50.0) | 9 (52.9) | 4 (44.4) |  |
| Asian/Pacific Islander | 1 (3.9) | 1 (5.9) | 0 (0.0) |  |
| Hispanic | 6 (23.1) | 2 (11.8) | 4 (44.4) |  |
| Other (e.g., decline to state,<br>multi-race) | 2 (7.7) | 2 (11.8) | 0 (0.0) |  |
| Education, n (%) |  |  |  | 0.95 |
| High school/GED or less | 2 (7.7) | 1 (5.9) | 1 (11.1) |  |
| College - 1 year to 3 years | 8 (30.8) | 5 (29.4) | 3 (33.3) |  |
| College - 4 years or more | 10 (38.5) | 7 (41.2) | 3 (33.3) |  |
| Post college | 6 (23.1) | 4 (23.5) | 2 (22.2) |  |
| Income, n (%) |  |  |  | 0.32 |
| < \$35,000 | 7 (26.9) | 4 (23.5) | 3 (33.3) | |
| \$35,000- <\$55,000 | 7 (26.9) | 3 (17.7) | 4 (44.4) | |
| \$55,000- <\$75,000 | 5 (19.2) | 4 (23.5) | 1 (11.1) | |
| >=\$75,000 | 7 (26.9) | 6 (35.3) | 1 (11.1) | |

Abbreviations: PST, problem-solving therapy.

### User Experience, Task Load, Working Alliance

Participants had a positive evaluation (values >0.8) for pragmatic (M ± SD, S1: 1.3 ± 1.2; S2: 1.4 ± 0.9), hedonic (S1: 1.0 ± 1.1; S2: 1.2 ± 1.0), and overall (S1: 1.2 ± 1.0; S2: 1.3 ± 0.8) qualities related to their user experience with Lumen for both sessions. There were no statistically significant differences between the two sessions (*t*(22)=0.37, 0.00, and 0.25, P values=0.71, 1.00, and 0.80, for pragmatic, hedonic and overall scores, respectively).

Across both sessions, participants encountered medium (∼50) across the mental (cognitive), effort, frustration, and the performance dimensions of the NASA-TLX scale, and there were no statistically significant differences between S1 and S2 (see Table 2). However, participants rated as having experienced more temporal workload in S2 (52.0 ± 29.1) than S1 (36.5 ± 23.2; P=0.03), suggesting feeling rushed during their interaction with Lumen in S2.

**Table 2.** Paired t-test Results Comparing NASA-TLX Scores between Sessions 1 and 2.

| Question | S1<br>(Mean±SD)<br>n=26 | S2<br>(Mean±SD)<br>n=23 | t value | P value |
| --- | --- | --- | --- | --- |
| How mentally demanding was the task?<br>(*mental demand*) | 42.7 ± 25.0 | 53.9 ± 26.1 | -1.80 | 0.09 |
| How hurried or rushed were you in the<br>pace of the task? (*temporal demand*) | 36.5 ± 23.2 | 52.0 ± 29.1 | -2.37 | 0.03 |
| How hard did you have to work to<br>accomplish your level of performance?<br>(*effort*) | 36.0 ± 23.4 | 42.8 ± 18.9 | -1.44 | 0.16 |
| How insecure, discouraged, irritated,<br>stressed, and annoyed were you?<br>(*frustration*) | 31.9 ± 22.0 | 38.5 ± 24.6 | -0.95 | 0.35 |
| How successful were you in<br>accomplishing what you were asked to<br>do? (*performance*) | 34.6 ± 23.1 | 37.2 ± 23.3 | -0.37 | 0.71 |

The scores on the 7-point WAI-tech survey for task (S1: 5.2 ± 0.9; S2: 5.3 ± 0.9), bond (S1: 4.9 ± 1.0; S2: 4.7 ± 1.0) and goal (S1: 5.0 ± 0.9; S2: 5.1 ± 0.9) subscales were moderately high, indicating that Lumen-based PST sessions were perceived to be aligned with the participants’ needs, addressing their potential goals and the degree of mutual liking. There were no statistically significant differences between both sessions on the task, goal, and bond scales or the overall scores (see Table 3).

**Table 3.** Paired t-test Results Comparing Task, Goal and Bond Subscales of WAI-T between Sessions 1 and 2.

| Scale | S1<br>(Mean ± SD)<br>n=26 | S2<br>(Mean ± SD)<br>n=23 | t value | P value |
| --- | --- | --- | --- | --- |
| Task subscale | 5.2 ± 0.9 | 5.3 ± 0.9 | 0.11 | 0.92 |
| Bond subscale | 4.9 ± 1.0 | 4.7 ± 1.0 | 1.49 | 0.15 |
| Goal subscale | 5.0 ± 0.9 | 5.1 ± 0.9 | -0.32 | 0.75 |
| **Overall Scale** | **5.0 ± 0.9** | **5.0 ± 0.9** | **0.56** | **0.58** |

### User Perspectives of Lumen

Based on the thematic analysis, we identified six categories that highlighted key user perspectives regarding Lumen. This included (% of each category across all transcripts): (1) comparing Lumen with a human coach (i.e., a human-AI comparison) (37%), (2) task load experienced during Lumen interactions (19%), (3) perception of PST delivered by Lumen (15%), (4) user suggestions for improving Lumen (15%), (5) natural language understanding of Lumen (8%), and (6) technical issues (5%) that were encountered during the two Lumen sessions (detailed descriptions of each of these categories along with exemplary quotations are provided in Table 4).

**Table 4.**
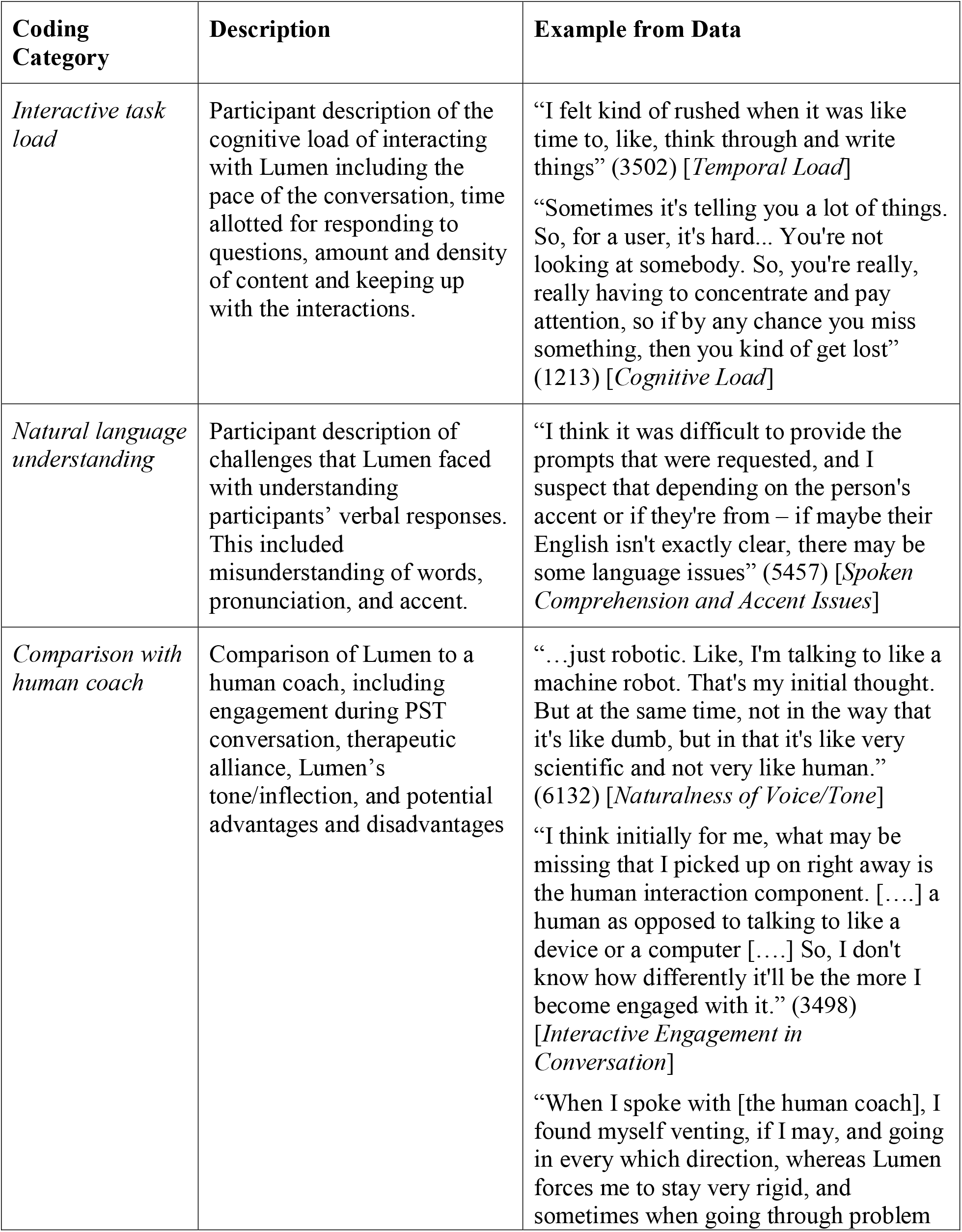
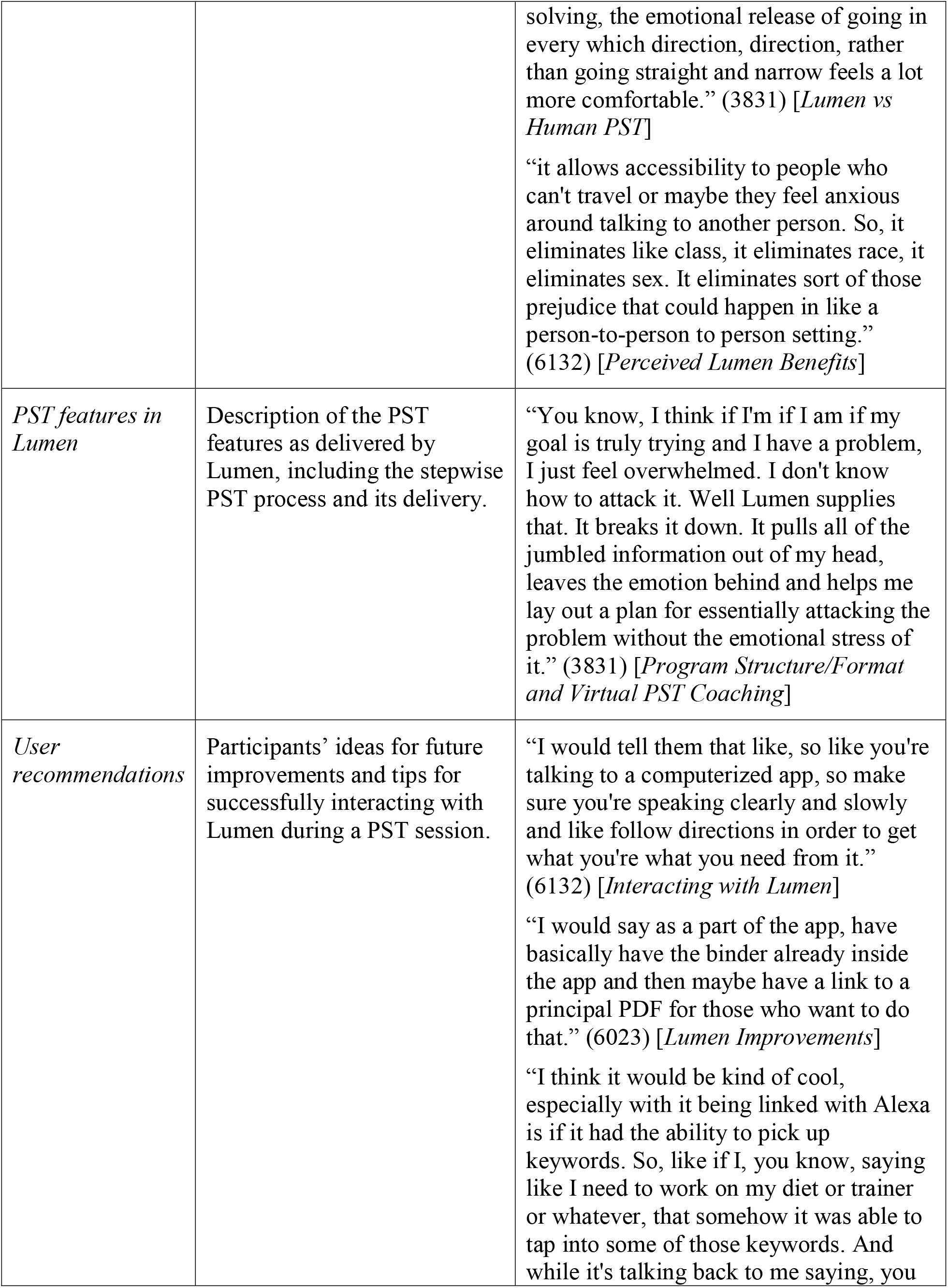
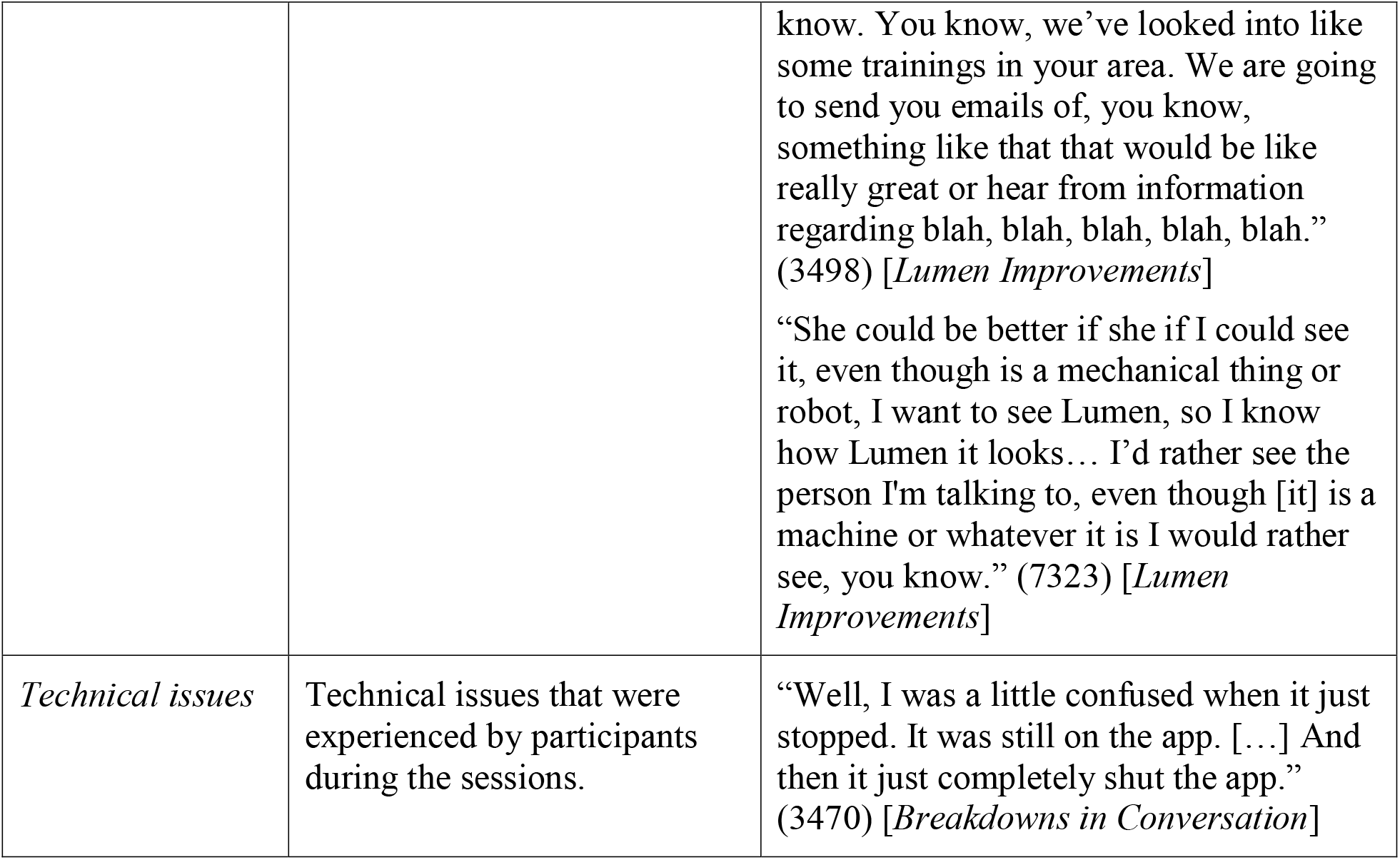
Coding Categories, Their Description and Examples from the Interviews.

| **Coding Category** | **Description** | **Example from Data** |
| --- | --- | --- |
| *Interactive task load* | Participant description of the cognitive load of interacting with Lumen including the pace of the conversation, time allotted for responding to questions, amount and density of content and keeping up with the interactions. | “I felt kind of rushed when it was like time to, like, think through and write things” (3502) [*Temporal Load*]<br>“Sometimes it's telling you a lot of things. So, for a user, it's hard... You're not looking at somebody. So, you're really, really having to concentrate and pay attention, so if by any chance you miss something, then you kind of get lost” (1213) [*Cognitive Load*] |
| *Natural language understanding* | Participant description of challenges that Lumen faced with understanding participants’ verbal responses. This included misunderstanding of words, pronunciation, and accent. | “I think it was difficult to provide the prompts that were requested, and I suspect that depending on the person's accent or if they're from – if maybe their English isn't exactly clear, there may be some language issues” (5457) [*Spoken Comprehension and Accent Issues*] |
| *Comparison with human coach* | Comparison of Lumen to a human coach, including engagement during PST conversation, therapeutic alliance, Lumen’s tone/inflection, and potential advantages and disadvantages | “...just robotic. Like, I'm talking to like a machine robot. That's my initial thought. But at the same time, not in the way that it's like dumb, but in that it's like very scientific and not very like human.” (6132) [*Naturalness of Voice/Tone*]<br>“I think initially for me, what may be missing that I picked up on right away is the human interaction component. [...] a human as opposed to talking to like a device or a computer [...] So, I don't know how differently it'll be the more I become engaged with it.” (3498) [*Interactive Engagement in Conversation*]<br>“When I spoke with [the human coach], I found myself venting, if I may, and going in every which direction, whereas Lumen forces me to stay very rigid, and sometimes when going through problem |
|  |  | solving, the emotional release of going in every which direction, direction, rather than going straight and narrow feels a lot more comfortable.” (3831) [*Lumen vs Human PST*]<br>“it allows accessibility to people who can't travel or maybe they feel anxious around talking to another person. So, it eliminates like class, it eliminates race, it eliminates sex. It eliminates sort of those prejudice that could happen in like a person-to-person to person setting.” (6132) [*Perceived Lumen Benefits*] |
| *PST features in Lumen* | Description of the PST features as delivered by Lumen, including the stepwise PST process and its delivery. | “You know, I think if I'm if I am if my goal is truly trying and I have a problem, I just feel overwhelmed. I don't know how to attack it. Well Lumen supplies that. It breaks it down. It pulls all of the jumbled information out of my head, leaves the emotion behind and helps me lay out a plan for essentially attacking the problem without the emotional stress of it.” (3831) [*Program Structure/Format and Virtual PST Coaching*] |
| *User recommendations* | Participants’ ideas for future improvements and tips for successfully interacting with Lumen during a PST session. | “I would tell them that like, so like you're talking to a computerized app, so make sure you're speaking clearly and slowly and like follow directions in order to get what you're what you need from it.” (6132) [*Interacting with Lumen*]<br>“I would say as a part of the app, have basically have the binder already inside the app and then maybe have a link to a principal PDF for those who want to do that.” (6023) [*Lumen Improvements*]<br>“I think it would be kind of cool, especially with it being linked with Alexa is if it had the ability to pick up keywords. So, like if I, you know, saying like I need to work on my diet or trainer or whatever, that somehow it was able to tap into some of those keywords. And while it's talking back to me saying, you |
|  |  | know. You know, we've looked into like some trainings in your area. We are going to send you emails of, you know, something like that that would be like really great or hear from information regarding blah, blah, blah, blah, blah." (3498) [*Lumen Improvements*]<br>"She could be better if she if I could see it, even though is a mechanical thing or robot, I want to see Lumen, so I know how Lumen it looks... I'd rather see the person I'm talking to, even though [it] is a machine or whatever it is I would rather see, you know." (7323) [*Lumen Improvements*] |
| *Technical issues* | Technical issues that were experienced by participants during the sessions. | "Well, I was a little confused when it just stopped. It was still on the app. [...] And then it just completely shut the app." (3470) [*Breakdowns in Conversation*] |

Comparisons of Lumen to a human coach included several aspects: potential flexibility, ease of accessibility of Lumen for those that cannot attend face-to-face appointments, and cost-related advantages. Participants also highlighted the non-human nature of the interaction describing the lack of changes in tone, emotion, instant feedback, and desiring a “*more personalized human touch*.” Nevertheless, nearly all participants described the potential advantages related to Lumen’s accessibility, allowing those in need for therapy easily access a coach at any time: “*the fact that the flexibility of it, the fact that I could be at home, where I could be in my car, or that, you know, I could take a moment and stop at work and go in a quiet room instead of having to, you know, actually go out and, you know, go to a building, find parking, all of the inconveniences that come with [face-to-face] appointments…”* Additionally, and importantly, participants with previous PST experience expressed that the Lumen sessions were similar to the human coach sessions that they had previously engaged in.

Participants also highlighted the workload associated with Lumen sessions, sometimes describing the difficulty in pausing sessions to collect thoughts as they worked through the steps of PST. This was especially the case in S2, where participants were required to brainstorm multiple solutions to a problem then list the pros/cons of each solution. The workload challenges identified were related to pacing of the sessions (temporal load) and the amount of information that was directed at the participants (cognitive load). One of the participants explained that the short time to respond made them “*feel pressured to come up with something …[*..*]. But she [Lumen] did ask if I needed more time, but when I was responding my answers, I [still] felt like it was a short time and I almost felt cut off*.*”*

Participants described their perceptions of the PST program/structure as well as Lumen’s role in delivering PST. Their comments highlighted the importance of the PST stepwise structured approach and Lumen’s PST coaching that enabled them to create goals that could have been overwhelming: “*If my goal is truly trying and I have a problem, I just feel overwhelmed. I don’t know how to attack it. Well Lumen supplies that. It breaks it down. It pulls all of the jumbled information out of my head, leaves the emotion behind and helps me lay out a plan for essentially attacking the problem without the emotional stress of it*.*”*

Participants provided several suggestions for improvement. This included further personalizing the PST sessions, creating embodied avatars for Lumen, incorporating a friendlier voice, and investigating ways for reducing the task load associated with the interactions. One of the most insightful aspects was several participants highlighting the importance of cognitive “offloading [24].” This was especially aligned with the need to reduce the cognitive load associated with conversational interactions, especially in a problem-solving session (S2), where participants had to identify and work through a problem, set a goal, identify and evaluate possible solutions, and then devise a structured action plan to address the problem. Participants also suggested the need for visualizing their tasks—either digital or paper-based—that would help in organizing their thought processes and saving the notes for future interactions, as highlighted in the following quote: “*If it would have a way in app, I mean, […] but like a way to help me, a way to help track for me what my progress is*.”

Although there were a few instances of technical issues where the participants’ verbal responses were not comprehended by Lumen due to issues related to accent or ambient noise, these issues were minimal and most users noted the ease of interaction, as described in the following quote: “*I was pretty much impressed with how easy was to use and, you know, it wasn’t intimidating at all*.”

## Discussion

We designed and developed a voice-based virtual coach, Lumen, that delivers an evidence-based PST program for depression and anxiety. To the best of our knowledge, Lumen is one of the first, fully integrated, voice-based virtual coach for delivering behavioral therapy. As opposed to prior research that has primarily used voice assistants in information seeking, Lumen delivers therapy, aligned with the goals and principles of an empirically validated PST program. In this developmental evaluation, participants found the Lumen virtual coach to have high pragmatic usability and user experience, with limited task load during interactions. Participants also highlighted the considerable advantages of Lumen including its on-demand accessibility to a virtual therapist, the delivery of a complex PST task with a simplistic structure and organization for achieving their therapy goals. Moreover, although the second session required increased input, interaction and effort from the sessions S1 and S2 did not exhibit marked differences in any of the considered measures except for temporal load (associated with the pace of the conversations), which was highlighted by the participants in their interviews. In addition, participants highlighted the lack of personalization and deep engagement in the conversation, and the relative lack of emotional engagement in the conversations.

In response to the participant suggestions, several design changes were incorporated. In order to reduce the temporal and cognitive load (i.e., reducing the pace of the conversations), we incorporated multiple functionalities within Lumen. First, we split longer conversations (especially in S1, where Lumen was providing overview) into multiple shorter conversations to reduce the mean length of conversations between Lumen and the participant. Such shorter lengths of (but more frequent) conversations allow more interactive turns and have been shown to improve the common ground and engagement between conversational partners [5, 25-28]. Second, we developed functionality that allowed participants to repeat, pause and resume conversations. This allows participants to ask Lumen to repeat if they could not keep up with the content or to pause conversations in situations where they need to take a break and then resume later. Finally, we also broadly slowed down the pace of the conversations to reduce demand.

Additionally, based on suggestions we also developed a workbook to accompany Lumen, available in physical and digital forms. The workbook includes content corresponding to PST and simple worksheets for taking notes and facilitates brainstorming problem-solving goals, developing and evaluating potential solutions, and creating action plans. Such a cognitive aid helps in externalizing the thought processes [24, 29, 30], and to create a record for follow up after the session. Recording and brainstorming with tools also affords cognitive benefits, especially with older adults, such as prospective memory regarding the goals and action plans that were created [31]. We also developed several features linked to Lumen to further integrate contextual aspects regarding the user including their current status and progress. For example, participants can track their progress by viewing their sessions completed and responses to PHQ-9 and GAD-7 surveys on a user dashboard. Similarly, responses on the PHQ-9 and GAD-7 surveys were integrated into the Lumen session and reviewed during the session to help participants monitor the level of their depressive and anxiety symptoms.

Finally, we also heeded several privacy and security considerations for pragmatic implementation and testing in a real-world setting within the context of a planned pilot randomized clinical trial. Towards this end, we will afford trial participants access to the Lumen skill within the Amazon Alexa app on a fully encrypted and locked down iPad, with timed exits for non-use. This allows for preventing accidental recording issues that have been reported around the use of voice-based smart devices (see additional details in Supplementary Material Section B).

In spite of these changes, several aspects of Lumen’s design and interaction are limited by current AI-based voice technology. In particular, the natural language understanding challenges of voice-based technology are well-documented [10]. These include difficulties in parsing tone, accent, and pronunciation in spoken language, creating breakdowns in conversation and making it functionally impossible to have a free-form, open ended conversation with these devices. Additionally, current technology is also not able to discern differences in emotion or other verbal cues that are easily interpreted in face-to-face human conversations [5]. With ongoing improvements in the technology, these challenges are likely to be mitigated over time, allowing for continued improvement of Lumen for optimized user experience.

This mixed-method formative research study has several limitations. The study was based on a small sample of users (n=26) who used Lumen in a relatively controlled environment. However, participants were engaged in two sessions and performed the Lumen interactions without external support. Only two sessions were evaluated with participants and as such, we cannot characterize participant experience with the entire 8-session PST program. However, sessions 2-8 mirror the S2 trialed in this study. It is likely that participants will become more comfortable with the Lumen interactions in the later sessions as they become more familiar.

Notwithstanding those technological and research limitations, the findings from the formative evaluation and the subsequent improvements in design and functionalities position Lumen to be a “minimum viable product” that is highly acceptable to participants, appears to veridically reflect PST content, and is ready for potential real-world, pilot testing. Recruitment has started for the pilot clinical trial (ClinicalTrials.gov, NCT# 04524104) in which 60 adults with mild to moderate depressive and/or anxiety symptoms are being randomized in a 2:1 ratio to the Lumen intervention or the waitlist control group and followed for 4 months. The objectives of the pilot trial are threefold: (1) to determine the feasibility and acceptability of the Lumen virtual coach for delivering the 8-session PST program; (2) to assess neural target engagement by comparing changes in amygdala and dorsal lateral prefrontal cortex in functional neuroimaging between the Lumen intervention and waitlist control groups; and (3) to examine the relationship between neural target engagement and changes in self-reported measures of mood, coping, and psychosocial functioning. The pilot trial will provide the preliminary data needed to accelerate the clinical and translational research on this novel digital psychotherapy and catalyze future development and definitive efficacy clinical trials.

## Conclusions

With a goal of overcoming the lack of empirical evidence for AI-based voice applications in behavioral therapy, we developed a voice-only virtual coach, Lumen, for delivering PST. The findings from the formative evaluation highlight feasibility, accessibility and favorable user experience. Suggestions for more natural conversations and better contextual support have resulted in an improved, minimally viable product. Lumen is being tested in a clinical trial to evaluate its neural mechanism of action and therapeutic potential in depression and anxiety. If successful, Lumen can be a viable voice-based therapist offering a realistic and cognitively plausible verbal interaction for personalized and accessible mental health care, filling a gap in traditional mental health services.

## Data Availability

This study involves qualitative and survey data only. With appropriate permissions from the institutions human subjects protections office, data may be available for sharing.

## Data Availability

The data from this study, in a de-identified form, will be available on request and with appropriate data use agreements.

## Author Contributions

TK, CRR, NW, JS, and JM conceived the study; TK, CRR, EK, NL, EK collected the data; TK, CRR, EK, NL and NW were involved in the preliminary analysis; all authors participated in the interpretation of the results, writing of the manuscript, and critical review. All authors gave final approval for submission.

## Footnotes

1 © Lumen™ 2019-2021, The Board of Trustees of the University of Illinois.

## References

1. Topol, E.J., High-performance medicine: the convergence of human and artificial intelligence. Nature medicine, 2019. 25(1): p. 44–56.

2. [cited 2021 April 1]; Available from: https://voicebot.ai/2019/07/09/new-data-on-voice-assistant-seo-is-a-wake-up-call-for-brands.

3. Steinhubl, S.R. and Topol, E.J., Now we’re talking: bringing a voice to digital medicine.The Lancet, 2018. 392(10148): p. 627.

4. Sezgin, E., Militello, L.K., Huang, Y., and Lin, S., A scoping review of patient-facing, behavioral health interventions with voice assistant technology targeting self-management and healthy lifestyle behaviors. Translational Behavioral Medicine, 2020. 10(3): p. 606–628.

5. Kannampallil, T., Smyth, J.M., Jones, S., Payne, P.R., and Ma, J., Cognitive plausibility in voice-based AI health counselors. NPJ digital medicine, 2020. 3(1): p. 1–4.

6. Boyd, M. and Wilson, N., Just ask Siri? A pilot study comparing smartphone digital assistants and laptop Google searches for smoking cessation advice. PLoS One, 2018. 13(3): p. e0194811.

7. Kocaballi, A.B., Quiroz, J.C., Rezazadegan, D., Berkovsky, S., Magrabi, F., Coiera, E., and Laranjo, L., Responses of conversational agents to health and lifestyle prompts: investigation of appropriateness and presentation structures. Journal of medical Internet research, 2020. 22(2): p. e15823.

8. Miner, A.S., Milstein, A., Schueller, S., Hegde, R., Mangurian, C., and Linos, E., Smartphone-based conversational agents and responses to questions about mental health, interpersonal violence, and physical health. JAMA internal medicine, 2016. 176(5): p. 619–625.

9. Nobles, A.L., Leas, E.C., Caputi, T.L., Zhu, S.-H., Strathdee, S.A., and Ayers, J.W., Responses to addiction help-seeking from Alexa, Siri, Google Assistant, Cortana, and Bixby intelligent virtual assistants. NPJ digital medicine, 2020. 3(1): p. 1–3.

10. Bickmore, T.W., Trinh, H., Olafsson, S., O’Leary, T.K., Asadi, R., Rickles, N.M., and Cruz, R., Patient and consumer safety risks when using conversational assistants for medical information: an observational study of Siri, Alexa, and Google Assistant. Journal of medical Internet research, 2018. 20(9): p. e11510.

11. Alagha, E.C. and Helbing, R.R., Evaluating the quality of voice assistants’ responses to consumer health questions about vaccines: an exploratory comparison of Alexa, Google Assistant and Siri. BMJ health & care informatics, 2019. 26(1): p. e100075.

12. Palanica, A., Thommandram, A., Lee, A., Li, M., and Fossat, Y., Do you understand the words that are comin outta my mouth? Voice assistant comprehension of medication names. NPJ digital medicine, 2019. 2(1): p. 1–6.

13. Wilson, N., MacDonald, E.J., Mansoor, O.D., and Morgan, J., In bed with Siri and Google Assistant: a comparison of sexual health advice. Bmj, 2017. 359.

14. Ismail, H.O., Moses, A.R., Tadrus, M., Mohamed, E.A., and Jones, L.S., Feasibility of Use of a Smart Speaker to Administer Snellen Visual Acuity Examinations in a Clinical Setting. JAMA network open, 2020. 3(8): p. e2013908–e2013908.

15. Cheng, A., Raghavaraju, V., Kanugo, J., Handrianto, Y.P., and Shang, Y. Development and evaluation of a healthy coping voice interface application using the Google home for elderly patients with type 2 diabetes. in 2018 15th IEEE Annual Consumer Communications & Networking Conference (CCNC). 2018. IEEE.

16. Li, J., Maharjan, B., Xie, B., and Tao, C., A Personalized Voice-Based Diet Assistant for Caregivers of Alzheimer Disease and Related Dementias: System Development and Validation. Journal of Medical Internet Research, 2020. 22(9): p. e19897.

17. Ma, J., Rosas, L.G., Lv, N., Xiao, L., Snowden, M.B., Venditti, E.M., Lewis, M.A., Goldhaber-Fiebert, J.D., and Lavori, P.W., Effect of integrated behavioral weight loss treatment and problem-solving therapy on body mass index and depressive symptoms among patients with obesity and depression: the RAINBOW randomized clinical trial. Jama, 2019. 321(9): p. 869–879.

18. Schrepp, M., Hinderks, A., and Thomaschewski, J., Design and Evaluation of a Short Version of the User Experience Questionnaire (UEQ-S). Ijimai, 2017. 4(6): p. 103–108.

19. Hart, S.G. and Staveland, L.E., Development of NASA-TLX (Task Load Index): Results of empirical and theoretical research, in Advances in psychology. 1988, Elsevier. p. 139–183.

20. Kiluk, B.D., Serafini, K., Frankforter, T., Nich, C., and Carroll, K.M., Only connect: the working alliance in computer-based cognitive behavioral therapy. Behaviour research and therapy, 2014. 63: p. 139–146.

21. Clarke, V. and Braun, V., Thematic analysis, in Encyclopedia of critical psychology. 2014, Springer. p. 1947–1952.

22. Crowe, M., Inder, M., and Porter, R., Conducting qualitative research in mental health: Thematic and content analyses. Australian & New Zealand Journal of Psychiatry, 2015. 49(7): p. 616–623.

23. User Experience Questionnaire. Available from: https://www.ueq-online.org.

24. Larkin, J.H. and Simon, H.A., Why a diagram is (sometimes) worth ten thousand words. Cognitive science, 1987. 11(1): p. 65–100.

25. Brennan, S.E. and Clark, H.H., Conceptual pacts and lexical choice in conversation. Journal of Experimental Psychology: Learning, Memory, Cognition, 1996. 22(6): p. 1482.

26. Brennan, S.E., Galati, A., and Kuhlen, A.K., Two minds, one dialog: Coordinating speaking and understanding, in Psychology of learning and motivation. 2010, Elsevier. p. 301–344.

27. Clark, H.H., Using language. 1996: Cambridge university press.

28. Clark, H.H. and Brennan, S.E., Grounding in communication. Perspectives on socially shared cognition, 1991. 13(1991): p. 127–149.

29. Scaife, M. and Rogers, Y., External cognition: how do graphical representations work? International journal of human-computer studies, 1996. 45(2): p. 185–213.

30. Zhang, J. and Norman, D.A., Representations in distributed cognitive tasks. Cognitive science, 1994. 18(1): p. 87–122.

31. Liu, L.L. and Park, D.C., Aging and medical adherence: the use of automatic processes to achieve effortful things. Psychology and aging, 2004. 19(2): p. 318.

