## Supplementary Materials for "Design and Formative Evaluation of a Voice-based Virtual Coach for Problem-Solving Treatment"

Supplementary Material

### Section A: Lumen Architecture

Lumen is a voice-only virtual coach that delivers Problem Solving Treatment (PST) to counsel participants with depression and/or anxiety using the 7-step problem solving process and the SSTA (stop, slow down, think and act) method of coping. Lumen conducts interactive conversations providing appropriate responses based on participant input. In order to deliver seamless interactions, we developed a robust and parsimonious architecture that combines PST, integrated components to provide context for Lumen conversations, data storage for the interactions, and a software infrastructure for providing security and privacy for these interactions.

Lumen architecture (see **Figure 1**) was developed with input and consultation from researchers (in computer science, medicine, health service researchers, and psychiatry), software developers, and human computer interaction experts. The architecture was developed on Amazon’s Alexa platform, with further integration with a secure REDCap system for ecological momentary assessments (EMAs) and surveys. Lumen’s software architecture is comprised of two components: a conversation manager module and a context manager module.

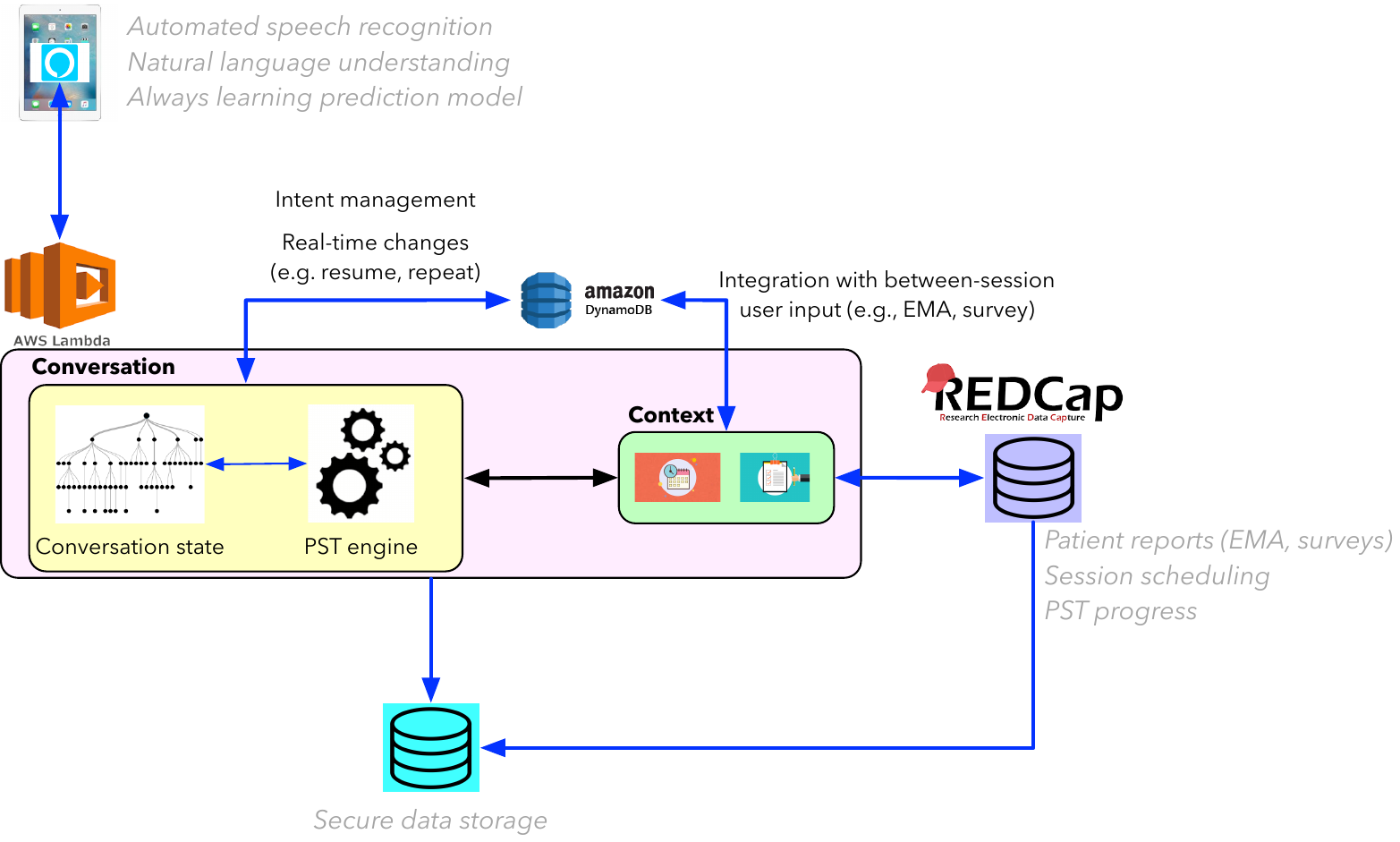

**Figure 1. Lumen software architecture that includes the conversation manager and context manager modules.**

The **conversation manager** is the voice-interactive component of Lumen and is responsible for delivering PST content. This PST content is aligned with the theoretical constructs and associated treatment guidelines. Towards this end, the conversation manager includes a PST engine that incorporates PST-related and conversational structures. The PST engine is a flexible set up that allows for the adaptation of PST content for other health-related problems (e.g., translating the therapy for weight management or smoking cessation) in the future.

Within the Lumen application, the PST engine tracks the progress of a PST session. For example, in a session, once the participant defines a specific problem to work on (step 1), Lumen will proceed to guide the participant in establishing a realistic goal for solving the problem (step 2). Then, once the participant has defined a set of potential solutions that can meet the goal (step 3), Lumen will guide the participant on evaluation of the pros and cons of each solution (step 4). After the participant chooses his/her preferred solution (step 5), Lumen will guide the participant to develop an action plan (step 6), which Lumen will prompt the participant to implement and evaluate the outcome post the session (step 7). This stepwise structure is the same in all PST sessions as is the case in current practice, and the participant identifies a problem to work on in each session (except session 1, which is an introductory overview session situating the PST process and building an initial problems list which participants can choose to work on in subsequent sessions, or they can opt to work on a new problem). Importantly, each of these steps is based on established PST theory and practice.

In addition, the conversation manager also manages the “state” of the conversation—including the flow of interactive conversation (e.g., the flow of the above-mentioned steps), and adaptive responses based on user input. The conversational state is adaptively managed based on user input. Towards this end, user “intents” or statements are parsed and mapped into pre-defined categories, which are then mapped to the appropriate state in the PST interaction to deliver relevant content, aligned with the user input. The conversation manager also provides dynamic support for functions such as resume, which allows participants to re-start a previously incomplete PST session. Such stop-restart functions provide flexibility in conversational interactions, affording perceptions of realistic conversational interactions.

The conversation manager also interacts with a **context manager** module that provides situated and contextual information regarding Lumen sessions. The context manager primarily controls three aspects: persistence of therapy content across sessions, scheduling/re-scheduling of sessions, and integrating external content (e.g., surveys, EMAs) into the Lumen sessions. The context manager dynamically tracks user identified problems (see step 1 above), and goals that were previously developed and asks participants to self-evaluate adherence to their previously developed action plans in ensuing sessions. Such dynamic follow-up increases the persistence and continuity of therapy across sessions, developing trust and confidence in the virtual coach. Similarly, the context manager tracks session progress (e.g., session 5), and helps participants schedule and re-schedule sessions. With its integration with an external scheduling database (in REDCap), follow-up emails and messages are tracked to ensure that the therapy sessions are synchronized with participant needs. Finally, the context manager also interacts with the REDCap database to incorporate survey responses (e.g., completion of the PHQ-9 and GAD-7 surveys before each session) into the Lumen session. For example, prior to the start of each Lumen session, participants are asked to complete the PHQ-9 and GAD-7 surveys; if incomplete (or partially complete), Coach Lumen prompts the user to complete the surveys then re-start the session.

All sessions and communicative interactions are stored in a secure AWS-based database for analysis. In order to prevent accidental recording, and for pragmatic implementation in a clinical trial, we currently have implemented the entire Lumen infrastructure in a “locked down” mode in an 8^th^ generation Apple iPad. Participants can access the Lumen application skill within the Alexa application, preloaded on their study iPad, by stating “Alexa, Open Lumen….” A summary of the Lumen components is provided in Table 1.

**Table 1. Lumen components, and their associated functions.**

| **Lumen Component** | **Functions** |
| --- | --- |
| Conversational manager | Managing conversations based on user intent, aligning with PST constructs, additional functions to manage conversations (e.g., repeat, resume), tracking progress within a session |
| Context manager | Tracking user problems and goals (from previous PST sessions), prompting participants to self-evaluate action plan adherence, integrating user responses from PHQ-9 and GAD-7 surveys within sessions, scheduling/re-scheduling sessions, |
| Lumen REDCap database | Ecological daily assessments, patient surveys, patient calendaring and reminders |
| Storage/Data Management (AWS & REDCap) | Comprehensive storage of user responses to surveys, user interactions with the Lumen, synchronization across devices, data security and privacy |

### Section B: Interacting with Lumen

Lumen architecture that is presented in Section A, is realized through user interactions with the Lumen skill embedded within the Alexa application (currently delivered on an iPad device) and user completion of surveys and EMAs with hyperlinks delivered via emails and text messages (on the participant mobile phones) (see **Figure 2**).

Participant interaction with the Lumen PST coach involves two primary components: (a) sessions with the coach, and (b) completion of surveys, EMAs, administrative management (e.g., session scheduling, re-scheduling, monitoring progress on their personal dashboard).

Participants access their scheduled Lumen session via the Alexa application on their assigned iPad. Sessions are instantiated by the participant by saying “Alexa, Open Lumen.” Conversations then continue based on the progress that participants have made (e.g., number of sessions completed), current problem(s) being addressed and implementation of previously created goals and action plans. As previously described, these conversations are aligned with PST’s treatment protocol and guidelines and with conversational structures and flow that reinforce the patient-centered approach of PST. As with regular conversations, participants can exit, resume or ask the Lumen coach to repeat parts of the conversations that they are unable to follow. At the end of each session, participants schedule their next session with Lumen.

Lumen’s PST sessions are delivered on a dedicated iPad, which is encrypted and functions in a “lockdown” mode with no additional functions or local storage. The purpose of using such an approach is for trialing it in a controlled environment without compromising on data safety and privacy of the user. Additionally, such an infrastructure and set-up provide several advantages. First, it creates the perception of a “device as a therapist” mode, assigning a specific role-based purpose (i.e., Lumen as a health coach, that is delivered only on the specific device). Second, embedding the Lumen skill within the Alexa application reduces the potential for accidental recording (as is common with smart speaker devices such as Echo or Google Assistant). Finally, the encrypted mode with no local data storage allows for data privacy protection and remote management, in case of a lost or misplaced device.

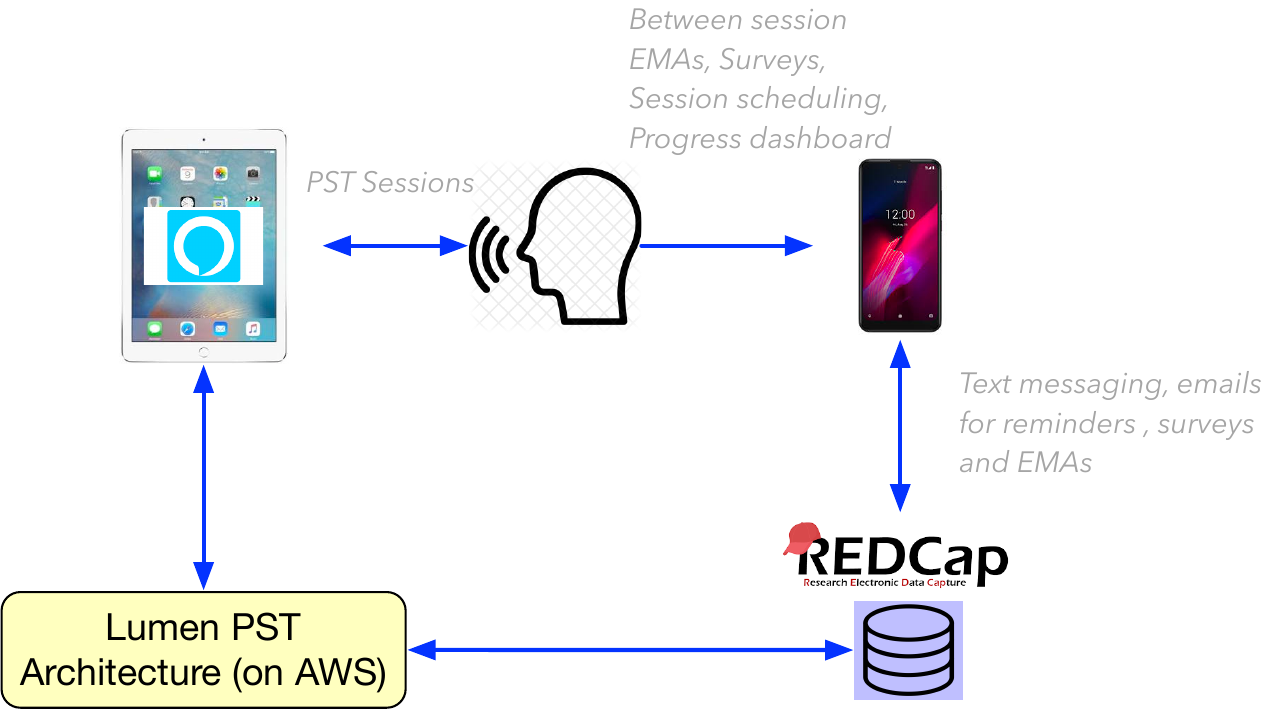

**Figure 2. User interaction with Lumen for PST sessions**

Between sessions, participants will also complete surveys and EMAs to track their progress. These administrative tasks and surveys, which provide situated context for Lumen interaction and monitoring of progress will be delivered on the participant’s personal devices. The surveys and EMAs are sent by text message with embedded links. Participants complete these on any browser associated with their mobile phone. Additional messages will include reminders about sessions, ability to schedule/re-schedule planned sessions, and other session related information.

All of the survey, EMA and scheduling tasks are instantiated through a dedicated REDCap project. Based on an initial session date, a program calendar according to PST guidelines (4 weekly, then 4 biweekly sessions) is set up and used for all session reminders. Future session appointments are confirmed or adjusted per user preference during Lumen sessions and are automatically updated in REDCap to assure that PHQ-9 and GAD-7 surveys are delivered at appropriate times.

### Section C: Components of PST Implemented in Lumen

To align with the treatment fidelity of the evidence-based PST, components of PST were implemented within the conversation manager and context manager of Lumen. The timing of the delivery of these components were aligned with the evidence-based PST. Each of the PST components, the timing of the delivery (i.e., session), and the aspects of the component managed by the conversation manager and context manager are provided in Table 1.

**Table 1. Components of PST that were implemented in various sessions within the conversation and context managers of Lumen, respectively.**

| **PST Fidelity Components** | **Session** | **Lumen Conversation Manager** | **Lumen Context Management (survey, EMA, participant workbook)** |
| --- | --- | --- | --- |
| **Introduction to Lumen and Building Rapport** | **Session 1** | Lumen introduces and tells participants about its role and purpose Personalized information | • Utilizes participant specific information stored in a profile |
| **Introduction to the Program and PST Overview** | **Session 1** | Natural conversation and verbal description of Lumen, background on PST and how it works |  |
| **Administer and Verify Completion of PHQ-9 and GAD-7** | **All Sessions** | Lumen checks and validates if surveys are completed within past 3 days, and, if not completed, provides directions on how to complete them | • REDCap links to complete PHQ-9/GAD-7 surveys externally sent via SMS. • Survey reminders sent before each session  • If session with Lumen begins and the PHQ-9/GAD-7 surveys are not complete, participant is asked to exit the session, complete surveys, then resume session • If PHQ-9/GAD-7 surveys are not completed within 3 days of completing Lumen session, participants sent new link and asked to re-take surveys. • REDCap links to complete brief (2-3 minute) EMAs that assess mood are sent for 7 days in a row, every other week. EMAs are open from 7PM to midnight on the day they are sent. If an EMA is not filled out by 9:30PM on the day it is sent, participants will receive one reminder to complete it. |
| **Knowledge Check** | **Session 1** | Multiple Choice questions asked by Lumen to evaluate participant understanding of key concepts before problem solving begins (e.g., link between unresolved problems and emotional distress, number of skills to be learned in the program, following a person-centered approach, rationale for utilizing PHQ-9/GAD-7, behavioral activation and planning varied activities) |  |
| **Problem List Generation** | **Session 1** | Lumen prompts participants to list problems they would like to work on in subsequent sessions | • Participants fill out the "Problem List" page in their workbook. |
| **7-Step PST Process** | **Sessions 2-8** | Lumen verbally walks participants through the steps of problem solving (Problem Identification, Goal Selection, Generating Possible Solutions, Solution Evaluation, Solution Choice, Action Plan Development). Lumen confirms (by repeating) participant answers and guides them to self-evaluate their responses at each step. Participants are asked if they need additional time as they complete each step.  Lumen will recall selected problems and goals and will ask participant about their action plan implementation at the next session as part of the progress review. | • Participants fill out "Problem Solving Worksheet" in their workbook to keep track of their listed problems. |
| **Progress Review** | **Sessions 3-8** | Prior to starting a new problem-solving session, Lumen asks participants to evaluate their action plan implementation from the immediate previous session  Lumen reminds participants of problems and goals they’ve chosen in the previous session. Further follow up is also conducted regarding how the participant was able to implement and adhere to the action plan, with Lumen prompting the participant to self-evaluate their action plan adherence. Additional questions regarding whether planned activities (Behavioral Activation) were carried out are also asked. | • Fill out "Progress Review" section of the workbook |
| **Behavioral Activation** | **Sessions 2-8** | Lumen explains 3 activities (social, physical, pleasant), their importance, along with examples. Participants are guided through planning 1 of each type of activities to partake in by the next session.  Lumen confirms participant activity choice and will ask participants whether they were able to complete their scheduled activities, at the next session, as part of the progress review | • Fill out "Behavioral Activation Worksheet" section of the workbook |
| **Scheduling, Rescheduling, Reminders** | **Between each session** | Participants are sent text message reminders about upcoming Lumen sessions (2/day) and asked to complete PHQ-9 and GAD-7 surveys before starting their upcoming Lumen session.  The timing of the sessions are based on session 1 (Lumen asks participant to write these down on the "Lumen Sessions" page in their workbook). Participants are encouraged to follow the pre-arranged session schedule; in case of emergency, participants have the ability to reschedule their Lumen session to a day before or after their pre-arranged session schedule. |  |
